# A Novel Method for the Estimation of a Dynamic Effective Reproduction Number (Dynamic-R) in the CoViD-19 Outbreak

**DOI:** 10.1101/2020.02.22.20023267

**Authors:** Yi Chen Chong

## Abstract

The CoViD-19 outbreak has escalated to a pandemic in the last few months. Pharmaceutical solutions based upon virologic studies, at this point, remain inconclusive, with no proven pharmaceutical solution so far. In contrast, this paper looks towards creating more accurate epidemiological models during this phase of viral growth in order to provide better feedback measures to public health officials and agencies, in particular, by providing a responsive, timely model of the R value based on the previous few days’ results.

Such an R value, although bearing less statistical precision due to limited sampling, could allow R to become a more effective, responsive standalone measure of infectious transmission. It demonstrates that the R value can be used as a dynamic, time-dependent indicator without the use of curve-fitting, and also estimates the most recent R-value of the CoViD-19 outbreak to be between 1.32 and 1.35.

## Introduction

The CoViD-19 outbreak has become increasingly severe, and no drugs so far have proved capable of stemming the viral growth. In contrast, this paper looks towards epidemiological models of the viral growth on a large scale. This paper attempts to provide a responsive, timely model of the effective reproductive number, R, which is used in many epidemiological models. This would allow governments to attempt to tackle the pandemic more effectively in terms of public health, as it provides a valuable and dynamic metric of the pandemic.

The more commonly known R_0_ value, or the basic reproduction number, measures how many secondary carriers an average infectious case will infect in an entirely susceptible population. The basic definition of R_0_ is the amount of infectious transmissions per unit time in contact with other people multiplied by the unit time in contact with others. Obviously, the R_0_ is dynamic. Public health measures aiming to reduce the transmissibility of the contact time people have with each other (e.g. curfews), or the number of infectious transmissions (e.g. through the use of protective equipment). The number has known to change during epidemics. For example, during the SARS outbreak, the R_0_ value dropped rapidly following public health measures that were taken. As such, the R_0_ value should be used as a dynamic indicator, and not necessarily as a stable predictor of future trends. The R value is an even more dynamic version of this. It is a measure of how many secondary carriers an average infectious case will infect, regardless of the susceptibility of the population. At the early stages of a pandemic, the 2 values are equal, but as the infected population begins to increase, and the susceptible population begins to decrease, the 2 values start to differ, and the R value begins to be a more valid indicator of the overall transmission of the disease.^1^

There are 2 main models of R estimation. The first model is to account for large amounts of data regarding behaviour of the infected population (e.g. traffic data), as well as data about the likelihood of transmission to build a simulation of predicted transmissions, usually with stochastic methods.^2^ This method is viewed as more accurate, as it factors in many different variables, but still has several flaws. First of all, the simulation obviously may not include confounding factors that have not been taken into account, causing inaccuracies. Secondly, it does not make sense to use R values to make predictions if said R values comes from predictions already. The R value is meant to be a simple indicator that can be used to make predictions. Third, the lack of responsiveness in the simulations means that the estimated R value is relatively static. If the R value changed rapidly, say, over the course of a few days, this model would be unable to keep up.

The second estimation method of the R value works with a simplified model of infection in 3 stages – Exposed, Infectious, and Isolated, which is a modified version of the SEIR structure. “Exposed” refers to the stage when a person becomes infected by a disease vector, but has not reached the stage where it can infect another person; “Infectious” refers to the stage when the person is a potential disease vector; “Isolated” refers to the stage where the person loses the ability to be a disease vector (i.e. the person either is isolated in quarantine, recovers, or passes away). This model was developed during the SARS epidemic by Lipsitch et al.^1^

This model develops a statistical approach – it discounts individual variables, using holistic statistics of cases per day, as well as basic viral data^1^. This vastly simplifies the problem, allowing for quick, responsive calculation. Furthermore, it uses R as a descriptor of past trends, rather than a predictor of future ones. Therefore, it is a more fitting method for my purpose of creating a responsive indicator of the current infectious situation.

## Method

The R estimator of this model deals with 3 variables. E and I are viral constants representing the lengths of the Exposed and Infectious stages respectively, in terms of time. K is the logarithmic growth rate, which is affected by many statistical factors, but bears the simple definition of being the rate of growth in the natural logarithm of the infected population. With these variables, R is defined as *R = K*^2^*EI + K*(*E* + 1) + 1.^1^

This model is also the one chosen by Cao et al in their preprint “Estimating the effective reproduction number of the 2019-nCoV in China”. E and I, as mentioned, are viral constants, and this paper will use the same values (E = 7 and I = 9). However, their methodology’s estimation of K is relatively static, and historical, using 6 timeframes over the course of the outbreak to create an average logarithmic growth factor.^3^ However, this method is prone to a couple flaws and insufficiencies. First, as a methodological error, using an average over 6 different timeframes of different length weights the K values in different time periods, producing a warped K-value, producing a final K-value closer toward the K-values during the shorter timeframes than an unweighted average. However, adopting timeframes of similar length would patch the flaw. Secondly, more paradigmatically, this method does not allow for the “dynamic indicator” approach to the R value. One key novelty, of the method presented in this work, therefore, will be in its dynamic estimation of the K value.

Similar models have also been used to try to find the instantaneous R values at any time t. Some models use regression to first perform curve-fitting on the dataset, before utilizing differentials of the curve equations to find the R values at any instance. This idea was first presented in Nishiura and Chowell’s “Fitting dynamic models to epidemic outbreaks with quantified uncertainty” and reused and refined in other works.^4,5^ However, this method presents several flaws. First of all, any changes in trends are not modelled by the curve fit. For example, if public health policies are implemented, and the rate of transmission drops significantly, a curve would smooth out that dramatic effect, and regard the sudden drop as an outlier. This removes a lot of the dynamism of the outputted model. In one particular model of the coronavirus outbreak, a preprint accounted for containment measures enacted on January 19, by creating a new trendline starting on January 29 (based on a 10.91-day incubation period).^6^ Manually correcting for error in such situations is not dynamic, and only accounts for changes that are known to have massive effect, and relies on a very precise knowledge of the incubation period. The method presented addresses that by only using recent data to calculate the R value on the day, which allows the R value to “respond” to drastic situations and changes. Secondly, a sufficient amount of data is required for curve fitting, which is not always available, especially near the start of an outbreak. The method presented in this paper only requires a few data points to make an estimation of the R value. Third, the method is exceedingly complex, with constant debate on the best methods of curve fitting, and the best models to use to calculate the R value. The method presented provides an exceedingly simple calculation method, that could be used even by those untrained in the studies of epidemiology or statistics.

As previously stated, K is defined to be the logarithmic growth rate, or 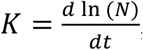 where N(t) is the number of confirmed cases at a certain time t (given in days from the start of the outbreak, which is 16 December, 2019, in the case of 2019-nCoV). Since the instantaneous growth rate is impossible to estimate without regressing to static regression-based curve-fitting models, it is obvious that the estimate has to rely on average growth rates. The process of an average logarithmic growth rate, therefore, is taking 2 different days, finding the difference in ln(N) between them, and dividing that value by the time elapsed. At first glance, it may seem impossible to “improve” the simple process. However, the selection of the days in of itself is a worthy problem. If the days you select are too far apart or too far in the past, the R value you generate loses its dynamism and currency; if you select too few days, you risk the chance of it being a statistical fluke.

In order for the data to be current, it should start from the most recently available data (most likely from the previous day). Furthermore, we should consider a range of K, with an average value, in order to clearly state the likely ranges of error. Therefore, I propose estimating the K value with the following function: 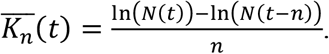.

In this set of functions, *n* represents how many days prior your sample is taken from (the larger the value of n, the more reliable your data is, but also the farther the sample size is in the past), and *t* represents the final day that your data ends upon. For the most recent indicator for the R value, for example, *t* would be set to the present day.

These functions could be better described this way. The average K value as of day *t*, from data collected over the prior *n* days, is found by dividing the difference in infected populations between the days by the number of days. The subscript and functional notation of R would remain the same as K, forming a set of 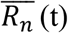. The findings of the R values could be stated as: in the n days prior to day t, an average virus carrier infected an average of 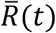 people. A confidence interval can also be constructed, and further analysis could be carried out with different values of n. However, the confidence interval constructed would most likely not have sufficient statistical data for small values of t.

A similar study was actually carried out last year in Japan, by Yamauchi et al, where the effective reproductive number of each week was calculated. In the definitions set out by this paper, their method calculates 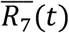, with 1-week intervals for t. However, their method does not provide a generalizable approach. Using different values of n could provide interesting, and more valuable, results. The method presented provides a notation, as well, for the estimation of the R value.

## Testing Methodology

In order to both test the method so as to generate relevant results for the current outbreak, as well as to prove the dynamism of the resulting R values, this paper uses the data from the European Center of Disease Prevention and Control^8^ on 2019-nCoV cases in the world to model the R(t) values over time, with both n=3 and n=5, over the period from January 5 (t = 20) until May 30 (t = 166).

## Findings

The graphs below show the computed values of 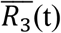 and 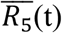, respectively, plotted over time. For those who would like to view the full data set, please contact the corresponding author.

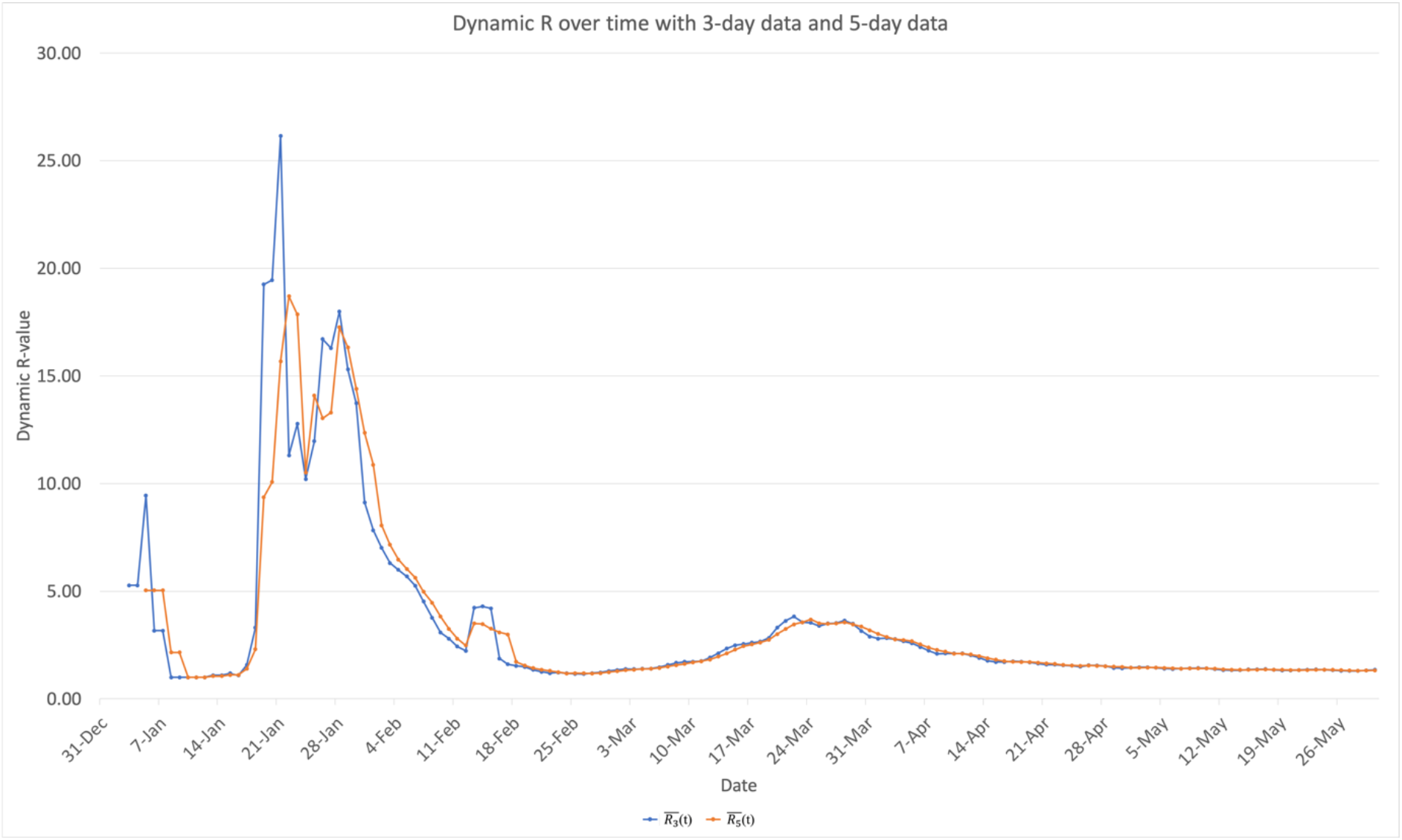

First of all, with regards to dynamism, the data more than demonstrates the dynamism of the R-value. There is massive change in the transmittivity of the data over time. The data shows a large spike in R(t) between January and February (near t = 42), followed by a sharp decline, and two more smaller spikes in a mostly downwards trend. A previous version of this paper analyzed the data in China between January and February, and detected a similar spike, suggesting that the spike could be due to factors including changes in the case reporting policy the prior week, the rising availability and ability of hospitals to carry out testing, etc. It also explains why some initial predictions of the infected population from before the end of January fall short of the current confirmed case count. The decline shows a relative positive outlook in that the transmission appears to be slowing down at least. However, generally, the R is also well above the previous Ro estimates of about 2-3 in the earlier periods. As for the most recent value, with the two estimates, it appears that the R value as of 30 May is between 1.32 and 1.35. A non-dynamic R-value calculated using all the data would provide an average of about 3.32, and no further information. A simple statistical confidence interval, however, in this case, would not reveal any valuable results, because, due to the small sample size, the range would be exceedingly large; furthermore, the K value is small and should always be positive, and so the curve will not be normal, but instead, skewed right.

## Conclusions

The R value is shown to be a very dynamic and sensitive statistic. As such, the previous methods of finding long term averages as if the value does not provide very valuable information. The use of R as a predictor is not incredibly useful as well; its dynamism seems to make it more well-suited as a descriptor of transmissibility, rather than a predictor of future transmission. It could be used as a metric to determine how effective certain public health policies are, but in predictive models, it may be too fundamentally unstable. This explains why many previous models using a static R as a predictive variable did not make accurate predictions. As such, static R values are often misleading, and do not provide a good prediction of the situation. Trend-wise, it appears that the coronavirus R-value is about 1.3 as of the end of May, and generally decreasing, although the fact that the R-value is still above 1 suggests that the viral transmission itself is still not in decline.

## Discussion

The data used, as for most epidemiological studies relies on the number of confirmed cases, which could be very different from the number of actual cases, due to factors such as variations in case reporting policies and abilities to test for the disease. However, once testing becomes commonplace and case reporting is more or less standardized, such confounding variables should be minimized. Furthermore, the R value may not be constant in different locations, and such, might not provide a global value. Further research is needed to study the confounding factors that go into this dynamic-R value. It is also important to note that this dynamic effective reproductive number should be used with interpretation and analysis; any changes or discrepancies in the data could be due to many factors, and a more in-depth analysis is needed to determine what the values or trends mean in terms of public health.

## Data Availability

All data used in the paper are publicly available on the Internet.

https://www.ecdc.europa.eu/en/publications-data/download-todays-data-geographic-distribution-covid-19-cases-worldwide

